# Optimal strategies for quarantine stopping in France – General expected patterns of strategies focusing on contact between age groups

**DOI:** 10.1101/2020.04.21.20073932

**Authors:** Benjamin Roche, Andres Garchitorena, David Roiz

**Author notes:** Equally contributed.

## Abstract

Due to the COVID-19 pandemic, many countries have implemented a complete lockdown of their population that may not be sustainable for long. To identify the best strategy to replace this full lockdown, sophisticated models that rely on mobility data have been developed. In this study, using the example of France as a case-study, we develop a simple model considering contacts between age classes to derive the general impact of partial lockdown strategies targeted at specific age groups. We found that epidemic suppression can only be achieved by targeting isolation of young and middle age groups with high efficiency. All other strategies tested result in a flatter epidemic curve, with outcomes in (e.g. mortality and health system over-capacity) dependent of the age groups targeted and the isolation efficiency. Targeting only the elderly can decrease the expected mortality burden, but in proportions lower than more integrative strategies involving several age groups. While not aiming to provide quantitative forecasts, our study shows the benefits and constraints of different partial lockdown strategies, which could help guide decision-making.

## Introduction

The COVID-19 pandemic is a major global crisis that has generated massive sanitary damages, economic costs and social disruptions. The primary local outbreak happened in Wuhan, China, and has since been dispersed to more than 200 countries. As of 21 April 2020, over 2,5 million cases and 170,000 deaths have been confirmed, with the epicenter in temperate areas (Europe and North America). This global pandemic calls for immediate control but, in the absence of vaccines or effective treatments, the only possible actions rely on changes in human behavior to reduce contact rates between people (i.e. social distancing). According to current estimates of the R0, the epidemic will cease to grow only when at least 66% of the population has acquired immunity against the virus^1^. Given the severity of the disease, especially for older age populations and vulnerable groups, the potential death toll associated with the epidemic has prompted governments to take unprecedented social distancing measures^1^. As a result, a third of the world population is currently under lockdown, and macroeconomic policies are being put in place to try to contain a global recession that is now inevitable, with G20 countries committing about 5 trillion UDS just to keep the economies afloat^2^.

The timing and duration of social distancing measures is critical^3^, and several measures must be combined to achieve high effectiveness^1^. Social distance measures have been implemented as part of two overall types of strategies^1^. First, suppression strategies aim to reduce the effective reproductive number (Re) to below 1, so that each new infection generates less than one other infection, progressively halting human-to-human transmission. The main challenge of this approach is that social distancing measures need to be effectively implemented, at more than 60 % of efficiency^4^, and also maintained for as long as the virus is circulating in the human population, or until a vaccine becomes available (at least 12-18 months). Suppression strategies are particularly effective in the early stages of the outbreak or when it is still locally confined, and have been successfully applied in China, South Korea, Japan, and Singapore^5^. Eventually, intermediate levels of local activity can be maintained while avoiding a large outbreak, as demonstrated in China and Hong Kong^6^.

Second, mitigation strategies aim to reduce and attenuate the health impact by building population immunity through the epidemic, leading to a more progressive increase and decline in transmission (i.e. flattening the epidemic curve), avoiding the saturation of the healthcare system^3^. Indeed, the sudden inflow of severe COVID-19 cases places an immense pressure on national health systems, with high demand for intensive care units (ICU) and mechanical ventilators rapidly outstripping capacity even in high-resource settings. Mitigation strategies across 11 European countries could already have averted 59,000 deaths (21,000-120,000 95% credible interval) in roughly three weeks^7^. Unfortunately, these interventions significantly delay reaching the threshold of immunity that is required for transmission to stop, stretching the economic impacts.

Mitigation measures in France have been progressively implemented, beginning with a ban of mass gatherings, school and university closures (March 13^th^-14^th^), social distancing encouraged and case isolation mandated (March 17^th^), leading to a full lockdown by March 18^th^ that will be in place at least until May 11^th^, 2020. Mitigation strategies in France and elsewhere can be implemented with different degrees of social distancing and target different groups, and could be optimized to minimize the number of hospitalizations and deaths while reducing their societal and economic impact^4^. The aim of this study is to evaluate under which conditions a partial lockdown strategy - where social distancing (quarantine) is targeted at specific age groups - could be an alternative to the complete lockdown currently in place in France, without an excessive impact on the expected epidemic burden.

## Materials and methods

### Mathematical model

We built a classic mathematical model in epidemiology based on a SEIR framework (Fig. 1). The population is divided into different age classes (characterized by the index *i*). Then, within each age class, the population is categorized according to their infectious status. First, individuals are in a Susceptible state (*S_i_*), where they can get the infection according to the basal transmission rate (*β*), the contact rate between age classes (*θ_ij_*) and the number of infectious individuals within each age class (*I_j_*). If infected, people move to the category Exposed (*E_i_*), where they are infected but not yet infectious. After an incubation period (*1/ε*, assumed here to be 3 days), individuals can become infectious and asymptomatic (*A_i_*) with a probability *p* (assumed here to be 80%), recovering after an infectious period (*1/σ*, assumed here to be 5 days) and then remain recovered for the rest of the outbreak (*U_i_*). We assume that asymptomatic individuals (because no symptom or a lack of detection) can transmit the virus like the symptomatic ones. Exposed individuals can also become infectious and symptomatic (*I*_*i*_) with a probability (*1-p*). In this case, they have a probability *α*_*i*_ (dependent of the age class) to develop a severe form of the disease (*M*_*i*_) and a probability *π*_*i*_ (also dependent of the age class) to die from the virus (*D*_*i*_). Otherwise, they recover and cannot be infected again (*R*_*i*_). *R*_*i*_ and *U*_*i*_ are identical from an infectious status, but the *R*_*i*_ will represent the cumulative incidence observed and reported in official sources.

**Figure 1:**
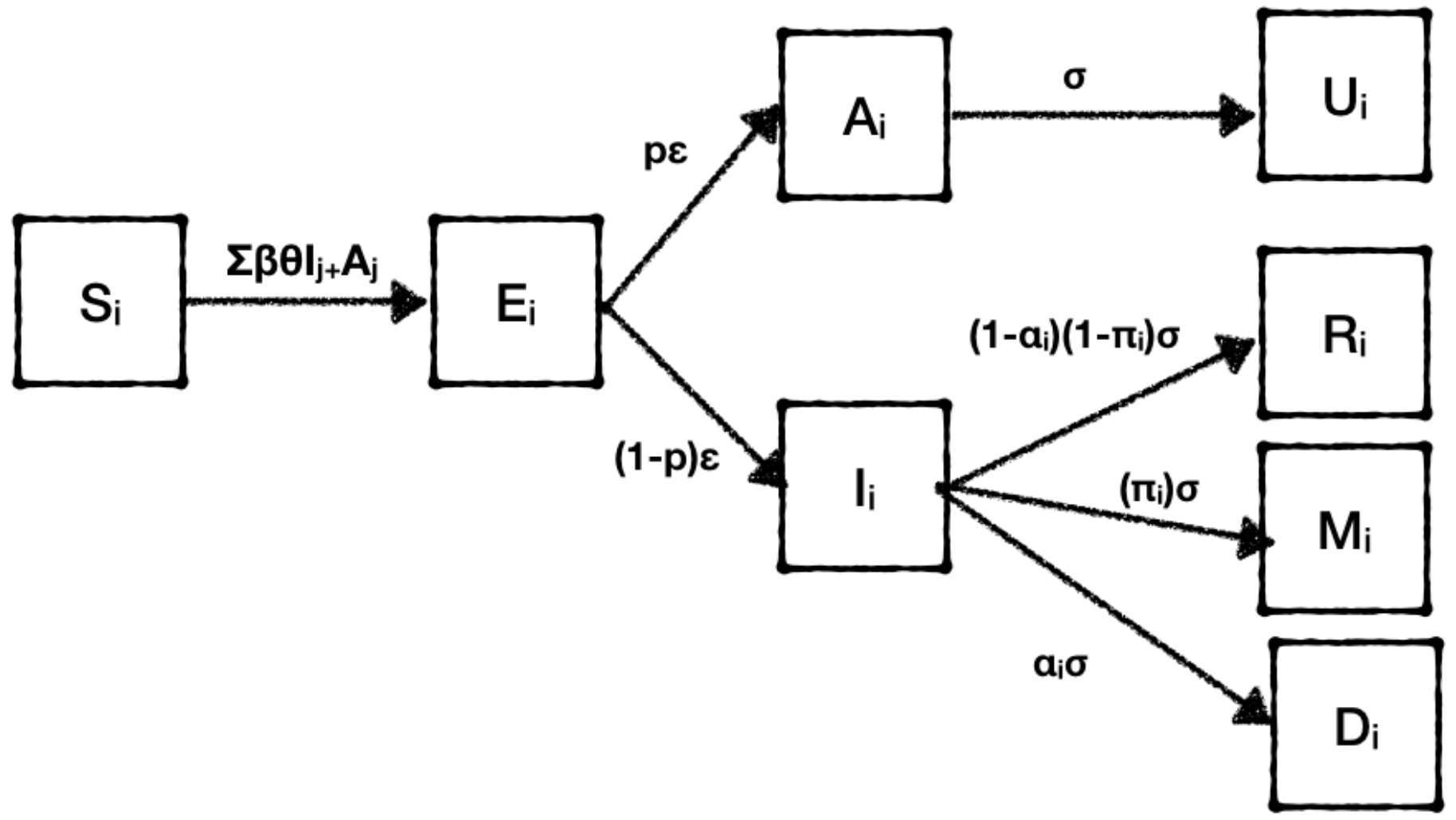
Structure of the mathematical model used for forecasting of COVID-19 in France, 2020.

We calibrated the model using the age structure of the French population from the National Institute of Statistics^8^, and contact rates between age groups from a large meta-analysis^9^ (Fig. 2). The severity per age group in terms of % of cases requiring critical care *α*_*i*_, and *π*_*i*_ the lethality of the considered age class (Infection Fatality Ratio) are detailed in Table 1 and are drawn from^1^.

**Table 1:**
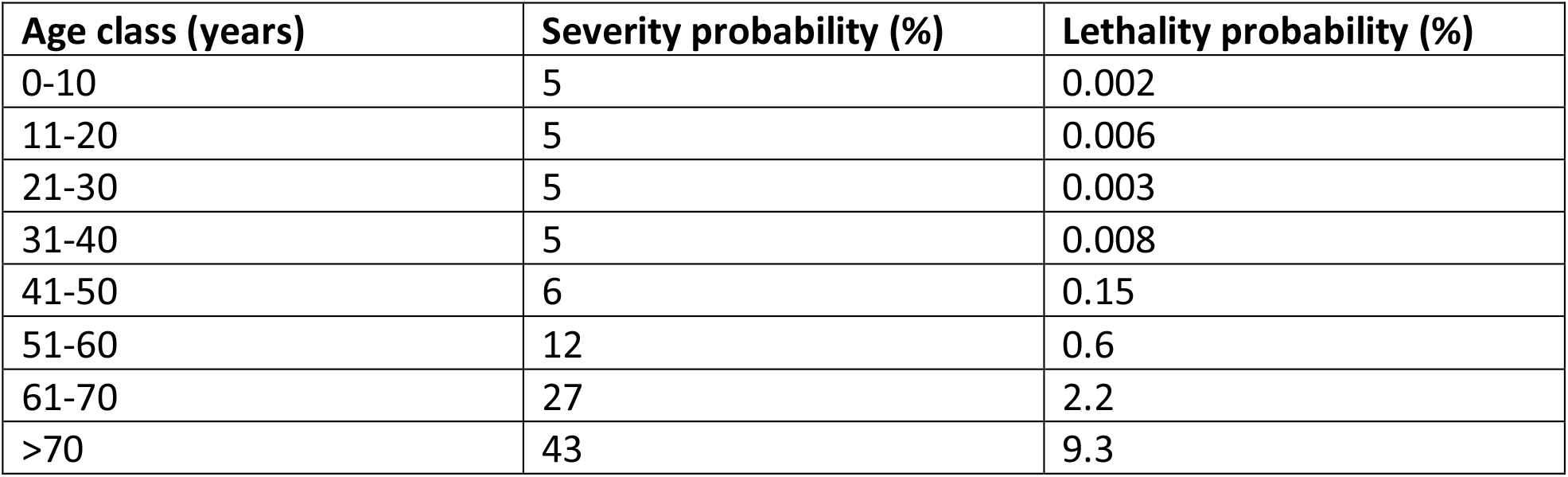
Severity and mortality probabilities^17^ by age group used in the simulations.

**Figure 2:**
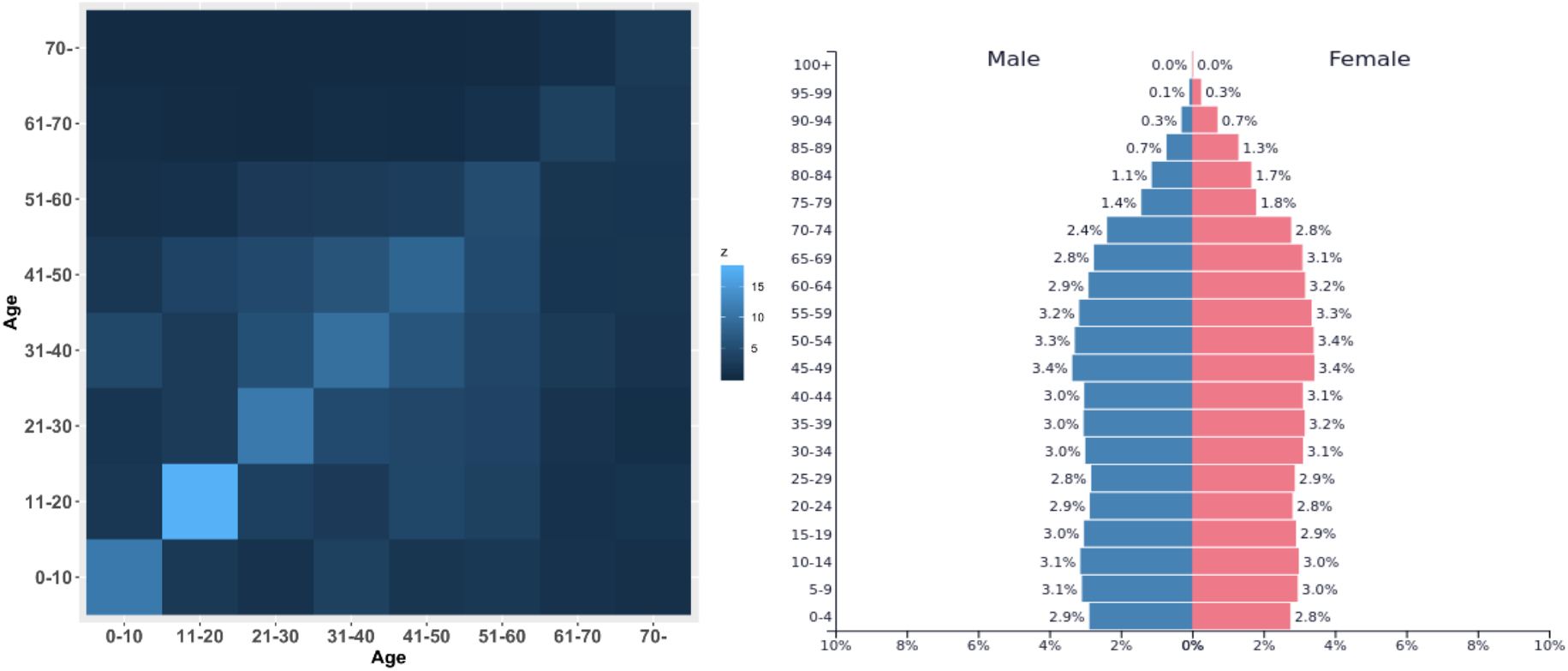
(Left) Matrix of age contact structure in France. Colours represent the number of daily contacts between each age class ^9^. (Right) Age pyramid considered for France^8^.

### Simulations and model analysis

We start the simulations on March 3^rd^, with 212 observed cases allocated uniformly within each age class. The initial number of non-observed cases is equal to the number of observed cases divided by the proportion of observed cases. We run our model with a transmission rate allowing a R0 of 2.8 and a proportion of non-observed cases at 80%, parameters that are consistent with other estimates^1^. We also assumed that the efficiency of social distance measures implemented on March 18^th^ was 50% until April 20^th7^.

After this date, we analyze scenarios with alternative social distance strategies to a strict lockdown as it has been implemented in France, starting in April 20^th^. With an estimation of the infectious status of the population at that time, we analyze three different strategies with their combinations (seven sub-strategies in total) that would impose a partial lockdown targeted at particular age groups (Fig. 3). The “Young age class”, assumed here to represent individuals between 0 and 30 years old, which could have decreased contact rates if both schools and universities are closed. The “Middle age class”, assumed here to represent individuals between 30 and 60 years old, could have lower contact rates if remote work (from home) is enforced. Finally, the “Elderly age class”, assumed here to represent individuals older than 60 years, could have less contacts if this age group is forced to stay home and retirement homes and other targeted places for this age group are isolated. We analyzed each of these strategies separately and in combination (Fig. 3). Each of the different partial lockdowns are evaluated according to their isolation efficiency (i.e. the efficiency to decrease contact rates of the targeted age group), which was varied between 0-100%. The different strategies are compared based on four outputs: the duration of the epidemic (assuming here that the epidemic could end when there is less than one new case per day), the cumulative number of deaths, the number of severe cases at the epidemic peak and the duration that the health system is overcapacity (assumed to be as 5,000 ICU beds).

**Figure 3:**
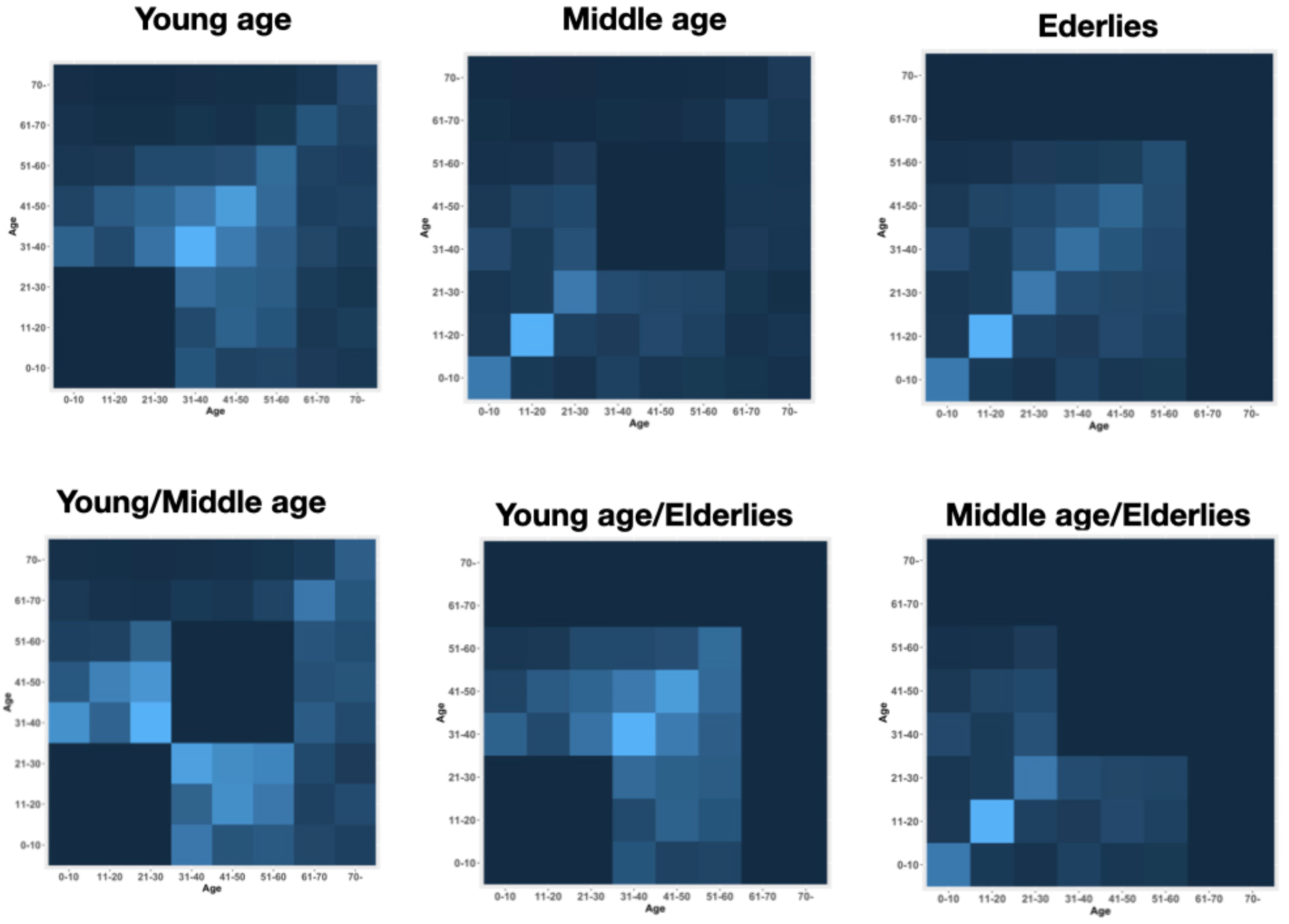
Assumed impact of the different partial lockdown strategies envisioned on the contact matrix between age classes. The colours represent the level of contact intensity, from low (dark) to high (light blue), with identical scale as in Figure 2. In this example, the isolation efficiency of each strategy is assumed to be 100%, so that the corresponding contact intensity for targeted age groups is zero.

## Results

We estimated that depending on the intervention implemented and its efficiency at reducing contact rates, the time before a halt in local transmission could range from less than 3 months to nearly two years (Fig. 4). The time estimated in the absence of interventions would be slightly under 1 year. In our models, a complete lockdown (i.e., targeting all age classes) and a partial strategy targeting young and middle age classes were the only interventions that could achieve suppression. This could be achieved at isolation efficiencies of at least 35% and 60% respectively, but their efficiency would have to be substantially higher in order to reduce the duration of the epidemic. Focusing only on elderlies did not dramatically change the epidemic duration regardless of the efficiency of this strategy in reducing contact rates. All other strategies flattened the curve, resulting on epidemic scenarios up to 200 days longer. Regarding the potential death toll, increases in efficiency for any strategy would consistently lead to reductions in the mortality burden, but these reductions were non-linear. A full lockdown was the most effective strategy at reducing mortality, resulting in about 9,000 additional deaths after April 20^th^ if efficiency is higher than 60%. Focusing on isolating the elderlies in addition to another age group (either young or middle age) was the next best strategy when the efficiency of interventions considered was lower than 50%, but the gap in mortality compared to a complete lockdown substantially increased after efficiencies over 20%. Targeting young and middle age groups could decrease indirectly the mortality in the elderly population and limit the total mortality burden to under 20,000 deaths when suppression was achieved promptly (efficiency higher than 75%).

**Figure 4:**
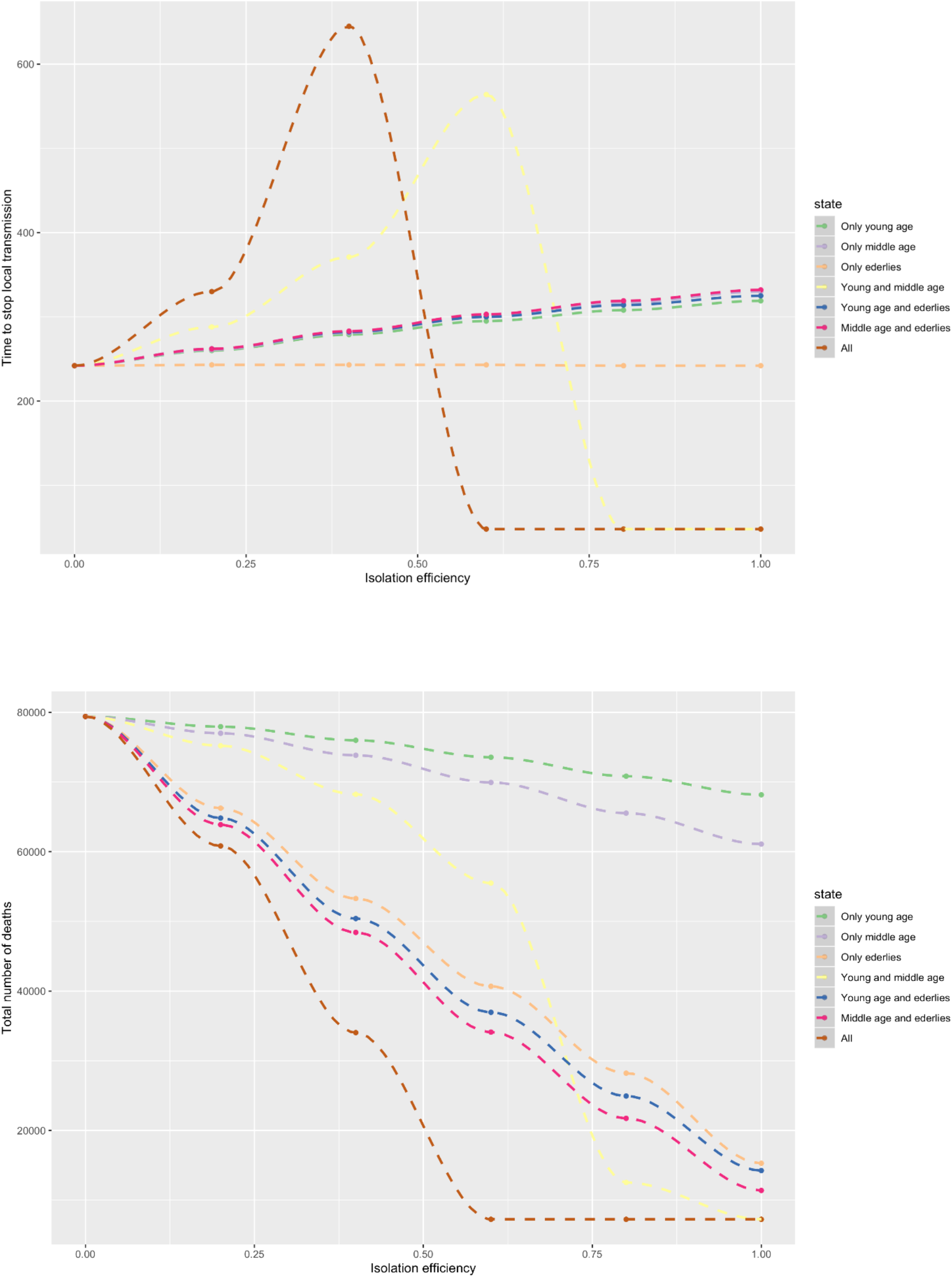
(Top) Time to break transmission and (Bottom) cumulative mortality for each of the seven different strategies according to their efficiency in decreasing contact rates with age classes targeted.

Similarly to mortality burdens, the ability of each strategy to limit over-capacity in the French health system improved at higher isolation efficiencies, but the effects were non-linear. In the absence of interventions, the health system would remain at over-capacity for 50 days, reaching over 20,000 severe cases at the peak. We found that a full lockdown remained the best strategy to limit this, avoiding over-capacity entirely at isolation efficiencies higher than 40% (Fig. 5). Focusing on young and middle age classes is an interesting lockdown strategy, halving the magnitude of the peak of number of severe cases while increasing the period in over-capacity at efficiencies lower than 40%, due to the shift between attenuation and suppression regime for this strategy around this threshold. Strategies including the elderly were less optimal but also limited overcapacity of the health system, leading to reductions of 50% in the length at over-capacity and of two thirds in the number of severe cases at the peak at efficiencies higher than 75%. Targeting only one age group resulted in considerable worse outcomes, compared to combined strategies, regardless of the age group targeted.

**Figure 5:**
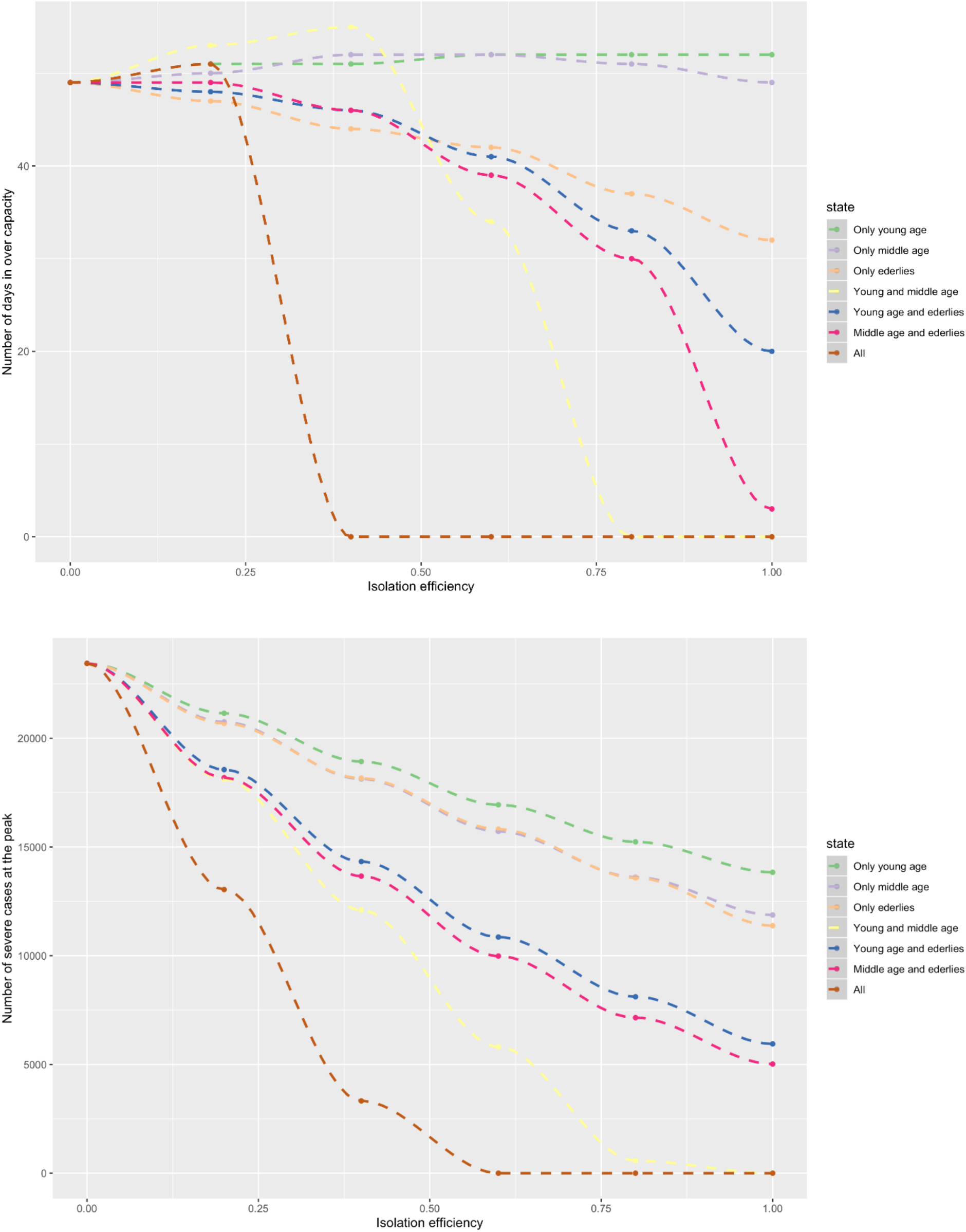
(Top) Number of days in health system over capacity. (Bottom) Number of severe cases at the epidemic peak. The outputs are for each of the seven different strategies according to their efficiency in decreasing contact rates with age classes targeted.

## Discussion

The ongoing COVID-19 pandemic has sparked worldwide a range of large-scale social distancing measures to a level not seen in over one hundred years^10^. The lessons learned from the 1918-19 Spanish flu epidemic highlighted the need to implement measures early and sustained over time to reduce overall mortality^10,11^. Unless herd immunity is achieved, transmission is expected to rebound following the lift of social distancing measures. In fact, to minimize mortality and health system overload, it is estimated that some form of social distancing measures will need to be in place for up to 18 months^1,12^, resulting in substantial impacts on the world’s economy^2,13^. In this study, we use an age-structured mathematical mode to simulate the impact of partial lockdowns - social distancing interventions targeted at specific age groups - in France as an alternative to maintaining a full lockdown. We show that while a full lockdown that achieves 60% isolation in the whole population remains the most effective strategy for minimizing both the morbidity and mortality burden of the epidemic (leading to suppression), targeting one or several age groups with higher efficiency could achieve comparable reductions while allowing important societal and economic activities to still take place.

The only partial lockdown strategy that achieved suppression in our simulations was targeting young and middle ages at efficiencies higher than 60%. The impact of this strategy on the outcomes assessed (epidemic duration, death burden, days over capacity and peak of severe cases) was comparable to a complete lockdown at an efficiency higher than 40%, because these age groups (young and middle ages) contribute greatly to transmission. However, at efficiencies around such thresholds, both these strategies could have important tradeoffs in the form of longer time to stop community transmission (up to nearly two years vs. one year with other strategies). This could involve substantial economic impacts, as the OECD estimates that each month of lockdown can lead to a loss of 2% in annual GDP growth^13^.

Since the majority of jobs even in western societies cannot be done remotely, a partial lockdown that avoids isolation of middle age groups would be preferable to reduce the economic impact of the epidemic. Among the strategies evaluated, we found that targeting the elderly and young age groups could lead to a large reduction in the number of deaths and minimize the impact on the health system when isolation efficiencies reach 80% or higher. Alternatively, targeting only the elderly population could theoretically decrease the death toll in similar proportions than if combined with another age group. This is consistent with other studies, where strategies involving isolation of the elderly showed the greatest potential to reduce mortality rates ^1^. However, relying just on targeting the elderly would result in thousands more severe cases at the peak of the epidemic than for other strategies, and the health system could be over capacity for several weeks. The consequences of this could be offset if current efforts to discover effective treatments and to rapidly increase the country’s bed capacity at intensive care units are successful. Yet, a health system over capacity for a sustained period could result in substantial all-cause excess mortality, something that has already been observed in several EU countries during the first weeks of the epidemic^3^.

This study has several limitations that deserve discussion. First, we used a deterministic model that does not consider the stochasticity in epidemic dynamics. Stochasticity is particularly relevant for predicting the tails of the epidemic curve, but its impact decreases as the number of cases increase (i.e. the bell of the curve). Thus, while it is unlikely to affect our conclusions, it may have resulted in an underestimation of the total epidemic time under different scenarios. Second, there is still uncertainty in the precise value of several parameters used in our model, and reports of confirmed COVID-19 cases and associated deaths are highly sensitive to testing rates and official case definitions. As a result, our assumptions may not hold true if there is a significant deviation in parameter values or between the real and reported burden of the epidemic. Third, we made multiple simplifying assumptions about age-specific contact rates, and we did not include transmission from pre-symptomatic cases. We used broad groups to roughly represent dynamics of populations whose primary activity is education, work or retirement so that we could envision age-specific measures for these groups, but in reality, these categories are fluid (e.g. individuals who start working at 18 years old, or who retire at 50). Moreover, we assumed that reducing contact rates within an age group (e.g. closing a school) would not impact contact rates with other groups (e.g. increased contact at home). This could have resulted in an overestimation of the impact of age-targeted interventions. Finally, there are many unknowns about the type and duration of the protective immunity for individuals after SARS-CoV-2 infection^14^.

The use of mathematical models has proved crucial for assessing the COVID-19 pandemic and the efforts to limit its consequences. As a result, an increasing number of modeling studies are arising, with a similar structure but varying degrees of complexity to allow a range of insights. At one side of the spectrum, Ferguson and collaborators adapted a formidable dataset and complex models their group had developed over decades to understand influenza dynamics ^15^. This allowed them to accurately estimate parameters for age groups, a variety of activities and locations, and explore in detail alternative measures to contain the COVID-19 epidemic in the UK and the US^1^. Yet, some of their main conclusions can be obtained from much simpler models, similar datasets are hard to obtain for other countries in a timely manner, and further complexity typically implies increased sensitivity to model structure and parameters. On the “simpler” side, models can accurately predict epidemic progression and provide several key insights with a minimal number of parameters and assumptions, but these are more limited in the range of questions that can be explored. Here, we leaned towards a simple model structure while keeping explicit consideration of age-specific compartments and their contact rates. Such information has now been estimated for over 150 countries from both developed and developing countries^9^. Therefore, while our model does not aim to forecast precisely the epidemiological dynamics at an accurate scale, it can be easily adapted to other settings and allow exploration of a wide range of scenarios and potential intervention measures.

Transmission models as the one presented here should be adapted to the particular context of developing countries to help understand potential epidemic impact and guide control efforts. Though the number of confirmed COVID-19 cases and deaths in developing countries remain low so far, there is still substantial uncertainty about how the epidemic will affect regions such as sub-Saharan Africa^16^. On one hand, a younger population structure in combination with lower connectivity due to poor road infrastructure could slow or limit the epidemic burden, especially in rural areas. On the other hand, high rates of malnutrition, respiratory infections due to indoor air pollution together with other comorbidities (e.g. infectious and parasitic diseases) could increase the severity of COVID-19 in younger ages due to impaired immunity or lung function. In addition, mortality rates of severe cases could be significantly higher than observed elsewhere due to lower rates of healthcare access and the limited capacity of health systems in the developing world (e.g. hospital bed capacity, availability of respirators, etc.). A full lockdown in these settings will be hard to sustain over time due to the dire economic conditions of populations and the limited resources of national governments to compensate households for the loss of revenue. Therefore, insights from such models could help find optimal alternatives adapted to the context of low-resource settings.

In conclusion, as social distancing measures are predicted to continue for several months, identifying the set of strategies that minimize the epidemic’s health burden with the least social disruption is essential in order to limit their collateral economic impact, both in developed and developing countries. Using the example of France, our results suggest that a full lockdown could be relaxed there without a substantial increase in the epidemic’s mortality burden if efficient age-targeted interventions are implemented. If a full lockdown is maintained in order to achieve suppression, it is critical that implementation of such measures achieve reductions over 50% in contact rates to avoid significant health and economic trade-offs. Similar studies could be adapted to other settings to allow countries make informed decisions about the best way to fight the epidemic.

## Data Availability

All data used are freely available

## Notes

### Competing Interest Statement

The authors have declared no competing interest.

### Funding Statement

This study has been partially funded by the ANR DigEpi

